# Long-term clinical, virological and immunological outcomes in patients hospitalized for COVID-19: antibody response predicts long COVID

**DOI:** 10.1101/2021.03.08.21253124

**Authors:** Javier García-Abellán, Sergio Padilla, Marta Fernández-González, José A. García, Vanesa Agulló, María Andreo, Sandra Ruiz, Antonio Galiana, Félix Gutiérrez, Mar Masiá

## Abstract

**Objective:** Long-term data following SARS-CoV-2 infection are limited. We aimed to characterize the medium and long-term clinical, virological, and immunological outcomes after hospitalization for COVID-19, and to identify predictors of long-COVID.

**Methods:** Prospective, longitudinal study conducted in COVID-19 patients confirmed by RT-PCR. Serial blood and nasopharyngeal samples (NPS) were obtained for measuring SARS-CoV-2 RNA and S-IgG/N-IgG antibodies during hospital stay, and at 1, 2 and 6 months post-discharge. Genome sequencing was performed where appropriate. Patients filled out a COVID19-symptom-questionnaire (CSQ) at 2-month and 6-month visits, and those with highest scores were characterized.

**Results:** Of 146 patients (60% male, median age 64 years) followed-up, 20.6% required hospital readmission and 5.5% died. At 2-months and 6-months, 9.6% and 7.8% patients, respectively, reported moderate/severe persistent symptoms. SARS-CoV-2 RT-PCR was positive in NPS in 11.8% (median Ct=38) and 3% (median Ct=36) patients at 2-months and 6-months, respectively, but no reinfections were demonstrated.

Antibody titers gradually waned, with seroreversion occurring at 6 months in 27 (27.6%) patients for N-IgG and in 6 (6%) for S-IgG. Adjusted 2-month predictors of the highest CSQ scores (OR [95%CI]) were lower peak S-IgG (0.80 [0.66-0.94]) and higher WHO-severity-score (2.57 [1.20-5.86]); 6-month predictors were lower peak S-IgG (0.89 [0.79-0.99]) and female sex (2.41 [1.20-4.82]); no association was found with prolonged viral shedding.

**Conclusions:** Late clinical events and persistent symptoms in the medium and long term occurred in a significant proportion of patients hospitalized for COVID-19. Gender, severity of illness and weaker antibody responses, but not viral shedding, were associated with long-COVID.

**Summary:** This study characterizes the long-term clinical, virological, and immunological outcomes following COVID-19 hospitalization. We found a significant proportion of late clinical events and persistent symptoms. Gender, severity of illness and weaker antibody responses, but not viral shedding, predicted long-COVID.

## INTRODUCTION

One year after the COVID-19 outbreak was first described [1], several questions about the disease remain to be answered. In contrast to the initial phases [2], long-term data following SARS-CoV-2 infection are limited. Dynamics of SARS-CoV-2 in the long-term, including the persistence of viral shedding, the incidence of late viral rebounds or reinfections, and their relationship with the clinical evolution of patients have not been defined. From the immunological perspective, another relevant question refers to the durability of the antibody response, and the impact of the intensity and duration of response on patients’ outcomes. In a significant proportion of patients, symptoms persist after hospital discharge for more than 2 months, which has been defined as long-COVID [3-5]. In addition to a more comprehensive characterization of the syndrome, the pathogenic mechanisms involved, including the role of viral shedding and the antibody kinetics, need to be determined. Acute respiratory distress induced by SARS-CoV-2 has been associated with persistent inflammation and pro-coagulation [6], which might potentially contribute to incomplete recovery, but the kinetics of inflammation and coagulation biomarkers after prolonged follow-up have not been disclosed.

We have longitudinally followed a cohort of patients hospitalized with COVID-19 who have been thoroughly investigated over a 6-month period after discharge. Our objective was to characterize the medium and long-term clinical, virological, and immunological outcomes, and to identify evolutionary trajectories and predictors of long-COVID.

## METHODS

### Study design, patients and study procedures

This prospective, longitudinal study was carried out at Hospital General Universitario de Elche, Spain. All patients admitted for COVID-19 between March 10^th^ and June 30^th^, 2020, were included in the analysis and were followed-up until 31^st^ December, 2020, the administrative censoring date of the study dataset. Cases included in the study were microbiologically confirmed through real-time polymerase chain reaction (RT-PCR) from nasopharyngeal swab samples in most cases and from fecal samples in 8.

Hospitalized COVID-19 patients were managed according to a predefined local protocol that included the diagnostic and therapeutic procedures during hospital stay [7]. This protocol consisted on the standardized collection of clinical variables and serial blood and nasopharyngeal sampling, obtained at different time-points during hospital stay for biochemical and sero-virological measurements.

Once discharged, patients’ follow-up was centralized at the Infectious Diseases Unit Outpatients’ clinic, where face-to-face visits were scheduled in the short (one month), medium (two months) and in the long-term (six months) after discharge. Phone and face-to-face visits not foreseen in the protocol were also appointed at the patients’ request.

At 2-month and 6-month visits, patients were offered to fill out a self-administered, self-rated COVID-19 symptom questionnaire (CSQ, Annex 1) in which they had to grade 11 items using a 10-point increasing intensity scale (0=absence of the symptom and 10=the maximum perceived intensity of the symptom). To characterize the persistence of symptoms, and as a test for robustness, we defined three different categories of the CSQ score: a score of at least one point (any symptom-CSQ); a score equal or more than the median (median-CSQ); a score above the third quartile (highest CSQ score) in any of the CSQ items. Scores were also classified into mild (1-4 points), moderate (5-7 points) and severe (8-10 points) categories.

In patients who missed scheduled appointments, an attempt was made to contact them by phone and their electronic medical records were carefully scrutinized to ensure the vital status of the patients. Hospital readmissions occurring during the 6-month follow up period were reviewed and recorded in the dataset.

### SARS-CoV-2 RNA and antibody measurements

RT-PCR analysis for SARS-CoV-2 was performed by means of a commercially available kit (AllplexTM 2019-nCoV Assay, Seegene, Seoul, Korea) which targeted the E, RdRP, and N genes. Suspected SARS-CoV-2 reinfection was defined according to the CDC criteria [8]. Genome sequencing of SARS-CoV-2 was performed on NPS samples following ARTIC amplicon sequencing protocol for MinIon version V3 (see Supplementary material for full description). IgG antibody plasma levels against the SARS-CoV-2 internal nucleocapsid (N) protein (N-IgG) (Anti-SARS-CoV-2-NCP IgG ELISA, Euroimmun, Lubeck, Germany) and surface S1 domain of the spike protein (S-IgG) (Anti-SARS-CoV-2 IgG ELISA, Euroimmun, Lubeck, Germany) were measured using commercial semi-quantitative EIA kits in an automated instrument (Dynex DS2® ELISA system). More procedures details can be found elsewhere [7].

### Statistical analyses

Continuous variables are expressed as median ± 25th and 75th percentiles (Q1, Q3), and categorical variables as percentages. Wilcoxon or Student's t-test were used to compare continuous variables, and the chi-square or Fisher's exact test for categorical variables comparison among patients with and without persistent symptoms. To compare the curves of viral load, antibody levels and biomarkers between groups, generalized additive mixed models were used. Interpolations in the graphs were carried out with cubic splines. Covariates with a p-value <0.05 in the crude comparison between groups and clinical relevant variables were included in multivariate analyses. Binomial logistic regression models were used to identify predictors of persistence of symptoms at 2 and 6 months. A receiver operating characteristic (ROC) curve analysis was performed to find the most discriminative serum level of anti-SARS-CoV-2 antibodies predicting persistence of symptoms. Statistical analysis was performed using R-project version 3.6.2.

## RESULTS

### Patient’s characteristics

A total of 162 patients were hospitalized with SARS-CoV-2 infection, 23 (14%) were admitted to the ICU and 12 (7.4%) died during hospital stay. After hospital discharge, 4 patients were lost to follow-up. Flow chart of patients is shown in Figure S-1. One hundred and four and 116 patients completed the SCQ at 2 and 6 months, respectively.

Baseline characteristics of the 146 included patients are shown in Table S-1. Median age was 64 years, 88 (60.3%) were male, and 72.6% had coexisting comorbid diseases. Clinical status on admission and therapy administered during hospital stay are detailed in Table S-1.

### Clinical and biological outcomes

During follow-up 30 patients (20%) were readmitted to hospital (Table S-2). The most frequent reasons for hospital readmission were underlying disease exacerbation (23 events in 13 patients, 8.9%), bacterial infection (12 in 5 patients, 3.4%), thrombohemorrhagic events (9 in 9, 6.1%), and persistent COVID-19 symptoms (8 in 6, 4.1%) patients. Eight (5.5%) patients died. Detailed information of causes of in-hospital and after-discharge deaths are shown in Table S-2.

Persistent symptoms of moderate or severe intensity at the 2-month visit were observed in 9.6%, 7.4% and 2.9% patients for general, gastrointestinal and respiratory symptoms, respectively, and in 7.8%, 4.3% and 1.0% patients, respectively, at the 6-month visit (Figure 1a).

**Figure 1.**
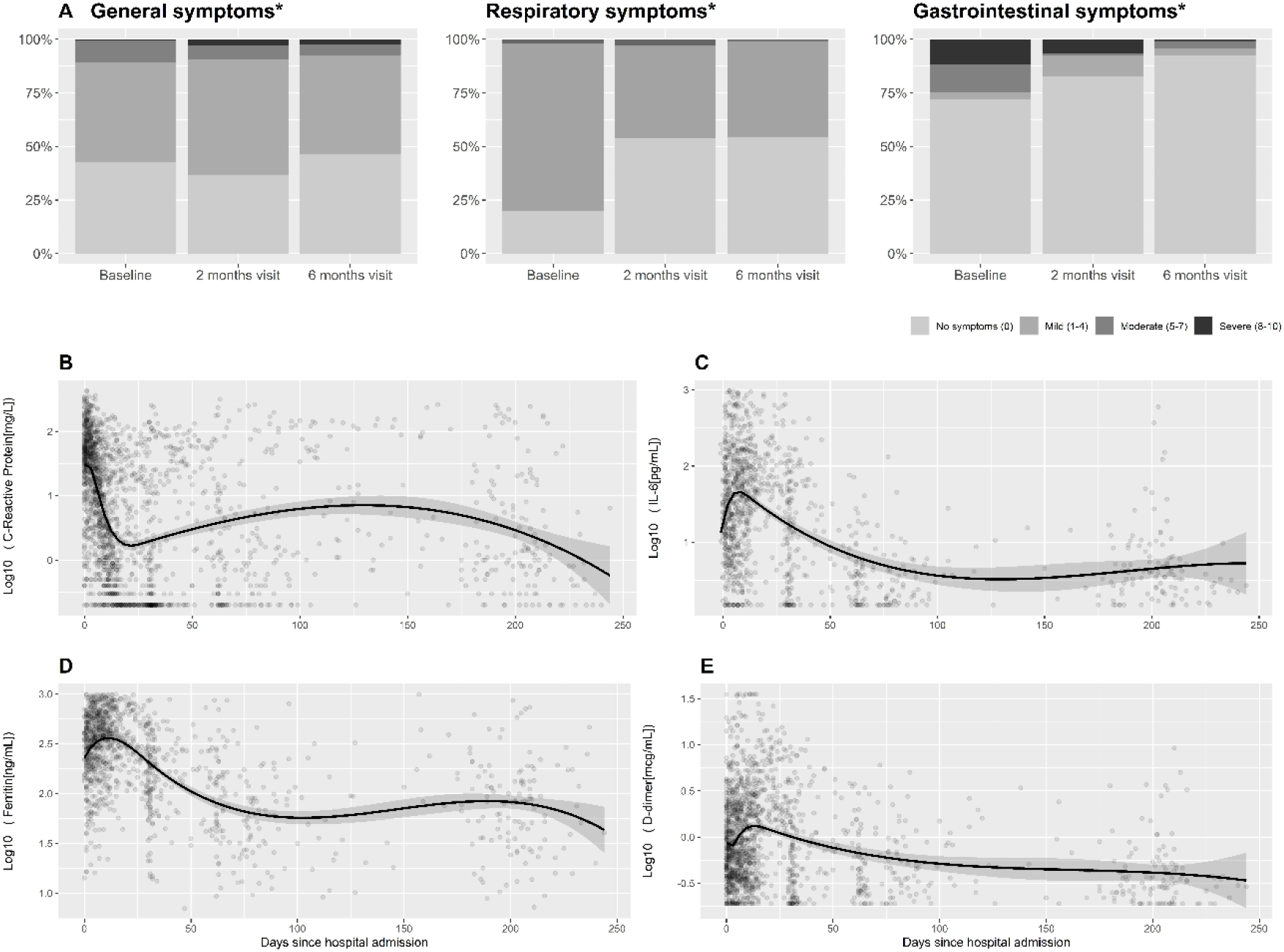
Temporal changes in symptom scores and serum levels of biomarkers during follow-up. Panel A shows the scores of self-reported general, respiratory and gastrointestinal symptoms included in the COVID-19 symptoms questionnaire at baseline, two months and six months. Panels B-E represent the temporal changes in the serum levels of C-reactive protein (Panel B), interleukin-6 (Panel C), ferritin (Panel D) and D-dimer (Panel E) since COVID-19 diagnosis. **Panel A:** General symptoms include fatigue, myalgia, sweating, headache; respiratory symptoms include cough, difficulty breathing, nasal congestion, sore throat, anosmia; gastrointestinal symptoms include diarrhea, vomiting, abdominal pain. *Pearson’s Chi-squared test *P-*value <0.05. **Panels B-E:** Each dot represents a serum biomarker value in an individual subject since hospital admission, with interpolation line and 90% confidence interval. Abbreviations: IL-6, interleukin-6.

Serum inflammatory biomarkers showed an initially steep and later flatter substantial decrease during follow-up, followed by stabilization or non-significant ulterior increase. The most prominent initial decrease was observed with C-reactive protein (CRP). Temporal changes in the levels of several biomarkers throughout the study period are shown in Figure 1b-1e.

### Virological outcomes

SARS-CoV-2 shedding lasted a median of 13 (2.2-33.8) days in those with the last RT-PCR test negative (Table S-1), resulting in a proportion of patients who tested negative at 2-month and 6-month follow-up visits of 88% and 97%, respectively. Viral shedding of low intensity was detected in some patients in subsequent nasopharyngeal tests beyond the acute phase of the disease. Thus, 40/146 (27%; median [Q1-Q3] Ct=34 [31-37]), 15/127 (11.8%; median Ct=38 [37.25-39]) and 4/134 (3.0%; median Ct=36 [36-36]) individuals tested positive at month 1, 2 and 6 visits, respectively. SARS-CoV-2 RT-PCR results during follow-up are displayed in Figure 2a. Despite no patient met the criteria for suspected reinfection [8], sequencing was performed in the 3 individuals with available paired samples of the 4 subjects with positive SARS-CoV-2 RNA at 6 months. The same clade 20B was present in all cases (Figure S-2). In two patients, the clade showed the same hallmark single nucleotide variants (individuals #88 and #95, Table S-3), and in the third patient, two new mutations were detected in the most recent sample (a K374R substitution in the N gene and an A222V substitution in the S gene), probably developed due to persistent infection.

**Figure 2.**
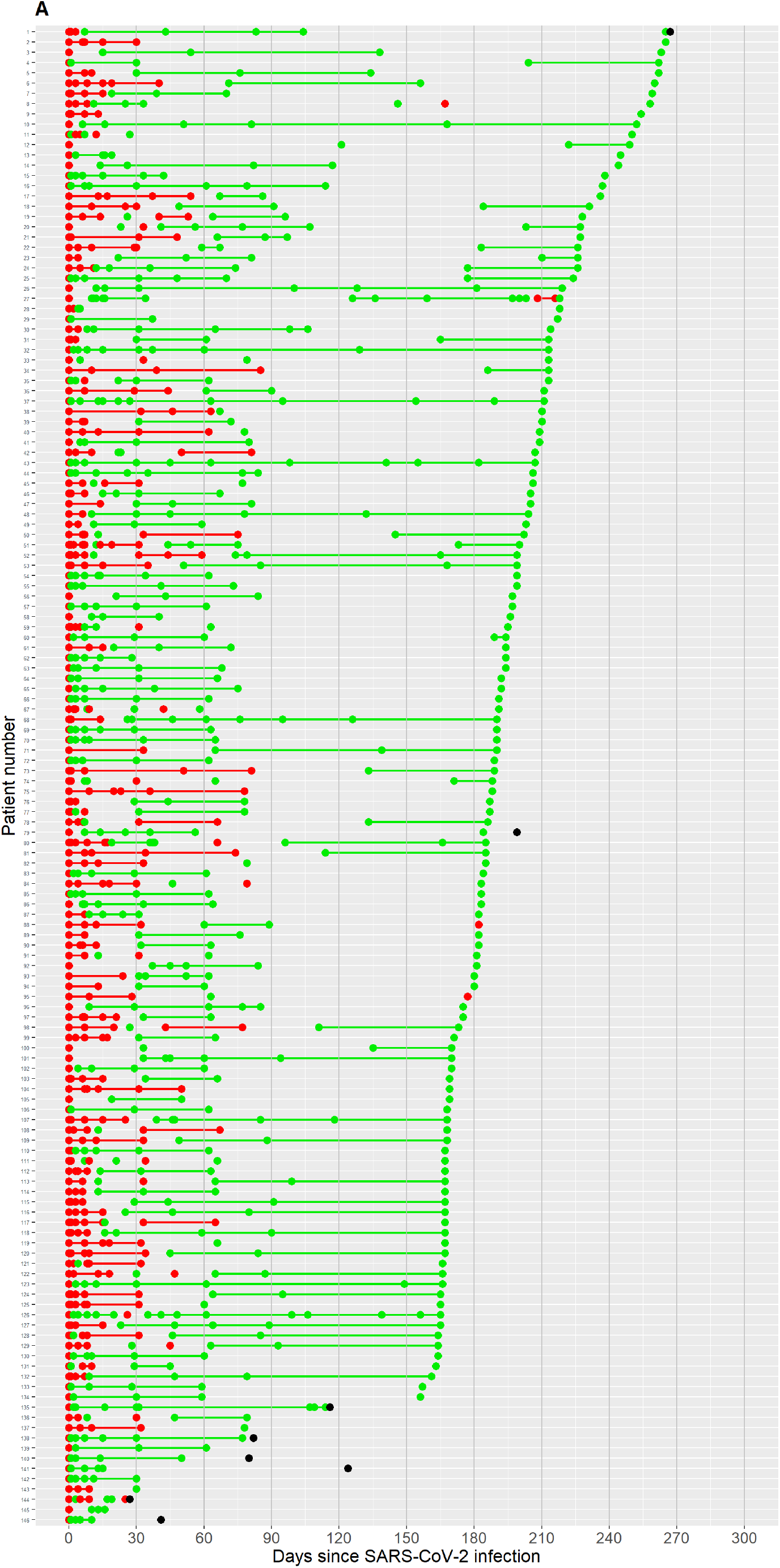
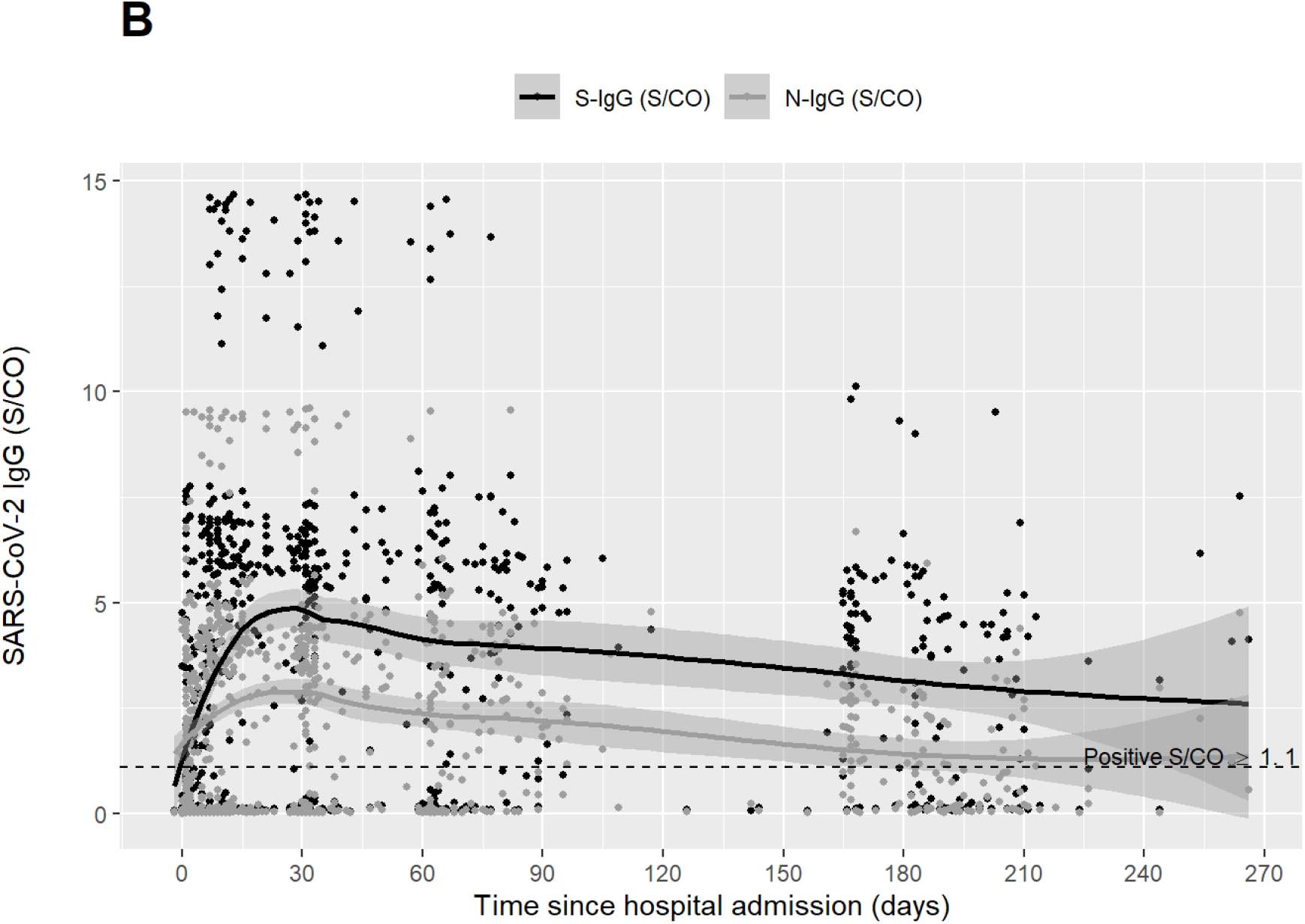
SARS-CoV-2 sero-virological changes during follow up. Panel A: SARS-CoV-2 RT-PCR test results during follow-up in the entire cohort. Panel B: Temporal changes in anti-SARS-CoV-2 surface S1 domain of the spike protein and nucleocapsid protein IgG antibodies. **Panel A:** Each horizontal level represents the RT-PCR SARS-CoV-2 results for each patient in the cohort throughout follow-up, since the first positive result to the last available one. Red dots represent positive results and green dots negative results. Black dots mark those patients who died and time of death since the first positive SARS-CoV-2 RT-PCR result. Colored lines join two consecutive dots in the same patient with the same result and less than 90 days between dots. Patient #143 moved to another country after discharge. Patients #139, #142 and #145 could only be reached by phone. Patients #136 and #137 refused to have a new nasopharyngeal sample taken but agreed to continue with the clinical follow-up. RT-PCR, reverse transcriptase-polymerase chain reaction. **Panel B:** Each dot represents an S-IgG (black) or N-IgG (grey) value in an individual subject, with interpolation line and 90% confidence interval Abbreviations: S/CO, absorbance/cut-off; S-IgG, S-IgG antibody against the SARS-CoV-2 surface S1 domain of the spike protein; N-IgG, antibody against the SARS-CoV-2 internal nucleocapsid protein.

### Serological outcomes

Median [Q1-Q3] time from illness onset to seropositivity was 12 (8-15) days. Peak S-IgG was significantly higher compared to N-IgG (median 5.9 vs 4.1 absorbance/cut-off [S/CO]; p<0.001, respectively) (Table S-1). Figure 2b shows S-IgG and N-IgG S/CO values of all determinations of the study patients over time. Antibody titers gradually waned, and 29 (28.7%) patients became seronegative for either N-IgG or S-IgG during follow-up: 23 for N-IgG alone, 2 for S-IgG alone, and 4 for both N-IgG and S-IgG. The majority of cases of seroreversion were observed at the 6-month visit (25/29 [86.2%] patients).

### Characterization and predictors of short-term persistence of symptoms

A significant number (76 [73%]) of patients scored any symptom-CSQ, being fatigue (54.8%), myalgia (30.8%), dyspnoea (26.9%) and cough (25%) the most frequent. The most frequent symptoms reported by patients with the highest CSQ scores were fatigue (12.5%), myalgia (7.6%) and dyspnoea (6.7%).

Table 1 shows the characteristics of the patients with the highest CSQ scores at 2 months. In the univariate analysis, they were more frequently active smokers (p=0.013), had more frequently bilateral lung infiltrates (p=0.015), exhibited lower baseline ferritin levels (p=0.002), a trend to lower CRP (p=0.055) and had received less frequently tocilizumab (p=0.013) during the acute episode of COVID-19.

**Table 1.** Clinical, sero-virological variables and serum biomarkers according to the persistence of symptoms at two and six months after hospital admission for COVID-19.

|  | Symptoms at 2 months |  |  |  |  |  | Symptoms at 6 months |  |  |  |  |  |
| --- | --- | --- | --- | --- | --- | --- | --- | --- | --- | --- | --- | --- |
|  | Yes* |  |  | No |  |  | Yes* |  |  | No |  |  |
|  |  |  |  |  |  | *P* value |  |  |  |  |  | *P* value |
| Patients, no. | 27 | (26) |  | 77 | (74) | - | 28 | (24) |  | 88 | (76) | - |
| Male sex | 12 | (44) |  | 51 | (66) | 0.066 | 14 | (50) |  | 55 | (62.5) | 0.274 |
| Age, years | 65 | (55-75) |  | 64 | (53-76) | 0.860 | 66 | (56-76) |  | 65 | (54-76) | 0.747 |
| Current smoking | 5 | (20) |  | 2 | (3) | 0.013 | 1 | (3.8) |  | 10 | (12.2) | 0.291 |
| **Comorbidity, no. (%)** |  |  |  |  |  |  |  |  |  |  |  |  |
| Any comorbidity# | 18 | (67) |  | 52 | (68) | 1.000 | 20 | (71.4) |  | 63 | (71.6) | 1.000 |
| CCI, median (Q1, Q3) points | 2 | (1-4) |  | 3 | (1-5) | 0.793 | 3 | (1-5) |  | 3 | (1-5) | 0.718 |
| Cardiovascular disease | 6 | (22.2) |  | 14 | (18.2) | 0.777 | 6 | (21.4) |  | 20 | (23) | 1.000 |
| Hypertension | 5 | (18.5) |  | 23 | (30) | 0.319 | 7 | (25) |  | 20 | (22.7) | 0.801 |
| Diabetes | 6 | (22.2) |  | 13 | (16.9) | 0.568 | 8 | (28.6) |  | 19 | (21.6) | 0.451 |
| Chronic obstructive lung disease | 1 | (3.7) |  | 6 | (8) | 0.674 | 3 | (10.7) |  | 5 | (5.7) | 0.398 |
| Symptoms duration, median (Q1, Q3) | 6 | (3-11) |  | 8 | (4-11) | 0.577 | 7.5 | (4.8-10) |  | 7 | (3-11) | 0.895 |
| No. of symptoms on admission | 3 | (2-4) |  | 3 | (2-4) | 0.654 | 3 | (2-5) |  | 3 | (1-4) | 0.425 |
| Maximum WHO severity score | 4 | (4-4) |  | 4 | (4-4) | 0.578 | 4 | (4-4) |  | 4 | (4-4) | 0.844 |
| SpO2/FIO2 on admission | 354 | (339-457) |  | 350 | (339-381) | 0.613 | 350 | (321-457) |  | 350 | (340-440) | 0.958 |
| Bilateral lung infiltrates in CR | 19 | (76) |  | 70 | (94.6) | 0.015 | 23 | (85) |  | 74 | (91.4) | 0.462 |
| Length of hospital stay, days | 10 | (7-14) |  | 11 | (8-14) | 0.494 | 11 | (7-14) |  | 11 | (7-16) | 0.354 |
| Admission to ICU | 5 | (18.5) |  | 6 | (7.8) | 0.148 | 3 | (10.7) |  | 12 | (13.6) | 1.000 |
| **Treatments** |  |  |  |  |  |  |  |  |  |  |  |  |
| Dexamethasone /Methylprednisolone | 3 | (11.1) |  | 14 | (18.2) | 0.549 | 4 | (14.3) |  | 16 | (18.2) | 0.778 |
| Tocilizumab | 9 | (33.3) |  | 48 | (62.3) | 0.013 | 15 | (53.6) |  | 43 | (48.9) | 0.829 |
| **Sero-virological features** |  |  |  |  |  |  |  |  |  |  |  |  |
| SARS-CoV-2 viral quantification | 1726 | (170-13869) |  | 4914 | (400-41608) | 0.262 | 1481 | (262-10379) |  | 2592 | (160-21023) | 0.900 |
| Time to viral clearance | 8 | (1-15) |  | 13 | (3.5-26.5) | 0.237 | 8 | (1-21.5) |  | 13 | (2-28) | 0.511 |
| RT-PCR SARS-CoV-2 positive, n (%) |  |  |  |  |  |  |  |  |  |  |  |  |
| ≥1 month | 5 | (18.5) |  | 27 | (35.1) | 0.174 | 10 | (35.7) |  | 24 | (27.3) | 0.466 |
| ≥2 months | 2 | (7.4) |  | 13 | (16.8) | 0.375 | 5 | (17.9) |  | 13 | (14.8) | 0.808 |
| Time to Seroconversion* | 16 | (11.5-28) |  | 11 | (8-13) | 0.012 | 12 | (9-16.8) |  | 12 | (8-15) | 0.413 |
| SARS-CoV-2 IgG-S |  |  |  |  |  |  |  |  |  |  |  |  |
| Seropositivity at 1 month, n (%) | 14 | (56) |  | 64 | (86.5) | 0.003 | 17 | (68) |  | 58 | (72.5) | 0.800 |
| Seropositivity at 2 month, n (%) | 15 | (55.6) |  | 65 | (86.7) | 0.002 | 17 | (68) |  | 61 | (72.6) | 0.801 |
| Seropositivity at 6 month, n (%) | 16 | (59.3) |  | 65 | (85.5) | 0.007 | 19 | (73.1) |  | 62 | (72.9) | 1.000 |
| Peak*, serum concentration (S/CO) | 4.1 | (0.1-6.2) |  | 6.4 | (4.5-7.5) | 0.002 | 5.1 | (2.2-6.1) |  | 6.3 | (2.6-7.2) | 0.055 |
| Trough* serum concentration (S/CO) | 3.5 | (2.0-4.5) |  | 3.8 | (2.4-4.7) | 0.346 | 3.9 | (2.0-4.8) |  | 3.7 | (2.2-4.7) | 0.987 |
| SARS-CoV-2 IgG-N |  |  |  |  |  |  |  |  |  |  |  |  |
| Seropositivity at 1 month, n (%) | 14 | (56) |  | 64 | (86.5) | 0.003 | 17 | (68) |  | 59 | (74) | 0.613 |
| Seropositivity at 2 month, n (%) | 15 | (55.6) |  | 65 | (86.7) | 0.002 | 17 | (68) |  | 61 | (72.6) | 0.801 |
| Seropositivity at 6 month, n (%) | 16 | (59.3) |  | 65 | (85.5) | 0.007 | 19 | (73) |  | 61 | (71.8) | 1.000 |
| Peak*, serum concentration (S/CO) | 3.6 | (0.1-5.0) |  | 4.3 | (3.5-4.9) | 0.210 | 3.8 | (1.8-4.7) |  | 4.1 | (2.6-4.8) | 0.563 |
| Trough* serum concentration (S/CO) | 2.3 | (1.9-2.7) |  | 2.0 | (1.4-2.6) | 0.171 | 1.7 | (1.3-2.6) |  | 2.2 | (1.5-2.7) | 0.124 |
| **Inflammatory biomarkers** |  |  |  |  |  |  |  |  |  |  |  |  |
| Serum C-reactive protein, mg/L |  |  |  |  |  |  |  |  |  |  |  |  |
| On admission | 25.3 | (3.5-59) |  | 49.2 | (23.2-104.3) | 0.055 | 21.6 | (2.5-45.4) |  | 47.5 | (15.3-130.6) | 0.014 |
| 1 month | 1.3 | (0.2-12.6) |  | 0.2 | (0.2-0.8) | 0.001 | 0.8 | (0.2-4.0) |  | 0.3 | (0.2-2.4) | 0.215 |
| 2 months | 1.7 | (0.7-10.9) |  | 0.3 | (0.2-1.0) | 0.001 | 1.0 | (0.3-4.2) |  | 0.4 | (0.2-4.8) | 0.357 |
| 6 months | 1.4 | (0.8-9.4) |  | 0.7 | (0.3-2) | 0.012 | 1.0 | (0.7-7.2) |  | 1.1 | (0.3-3.1) | 0.227 |
| Serum IL-6, pg/mL |  |  |  |  |  |  |  |  |  |  |  |  |
| On admission | 15.1 | (4.9-72) |  | 27 | (12-91.3) | 0.193 | 13.5 | (7.5-47.5) |  | 28.3 | (11.4-94.6) | 0.099 |
| 1 month | 7.7 | (3.4-52.5) |  | 19.9 | (3.4-49.3) | 0.682 | 18.1 | (3.5-75.6) |  | 13.8 | (3.3-48) | 0.504 |
| 2 months | 4.6 | (2.4-12.1) |  | 5.5 | (2.6-23.8) | 0.450 | 5.1 | (2.5-21.0) |  | 5.8 | (2.5-22.6) | 0.820 |
| 6 months | 3.7 | (2.2-4.7) |  | 2.9 | (1.8-5.1) | 0.454 | 3.6 | (1.5-6.8) |  | 3.1 | (1.8-5.2) | 0.965 |
| Serum Ferritin, ng/mL |  |  |  |  |  |  |  |  |  |  |  |  |
| On admission | 164 | (85-462) |  | 398 | (235-589) | 0.002 | 193 | (127-498) |  | 358 | (205-597) | 0.132 |
| 1 month | 129 | (60-311) |  | 181 | (98-291) | 0.219 | 100 | (53-256) |  | 195 | (106-364) | 0.020 |
| 2 months | 66 | (50.4-182.2) |  | 126 | (63-199) | 0.294 | 58 | (37-130) |  | 140 | (68-213) | 0.008 |
| 6 months | 57 | (19-109) |  | 77 | (38-129) | 0.233 | 53 | (23-88) |  | 76 | (32-130) | 0.239 |
| Serum D-dimer, mcg/mL |  |  |  |  |  |  |  |  |  |  |  |  |
| On admission | 0.7 | (0.4-1.5) |  | 0.6 | (0.4-1.2) | 0.784 | 0.7 | (0.4-1.3) |  | 0.8 | (0.4-1.9) | 0.656 |
| 1 month | 0.7 | (0.3-1.7) |  | 0.4 | (0.2-0.8) | 0.071 | 0.4 | (0.3-0.8) |  | 0.6 | (0.2-1.4) | 0.764 |
| 2 months | 0.6 | (0.3-1.4) |  | 0.4 | (0.2-0.7) | 0.069 | 0.5 | (0.3-0.8) |  | 0.5 | (0.2-0.9) | 0.746 |
| 6 months | 0.4 | (0.3-0.7) |  | 0.4 | (0.2-0.7) | 0.378 | 0.4 | (0.3-0.8) |  | 0.3 | (0.2-0.7) | 0.160 |
| NLR |  |  |  |  |  |  |  |  |  |  |  |  |
| On admission | 3.5 | (2.7-6.5) |  | 5.0 | (2.9-6.7) | 0.536 | 3.6 | (2.9-5.3) |  | 5.0 | (3.1-6.6) | 0.076 |
| 1 month | 1.8 | (1.3-2.2) |  | 1.7 | (1.3-2.4) | 0.758 | 2.0 | (1.5-2.8) |  | 1.7 | (1.1-2.6) | 0.163 |
| 2 months | 2.0 | (1.5-2.6) |  | 1.7 | (1.3-2.3) | 0.326 | 2.2 | (1.3-2.8) |  | 1.8 | (1.3-2.4) | 0.310 |
| 6 months | 2.1 | (1.7-3.2) |  | 1.9 | (1.5-2.3) | 0.160 | 2.0 | (1.8-2.6) |  | 2.0 | (1.4-2.7) | 0.338 |

\*Patients were allocated in the symptomatic group if the score on any of the symptoms in CSQ was in the top quartile of these symptoms' group scores. #This category included at least one of the following: diabetes, cardiovascular (including hypertension) respiratory, kidney, neurological disease, cirrhosis or malignant neoplasm. \*From illness onset and considering either S-IgG or N-IgG. CSQ, Covid-19 symptoms questionnaire; CCI, Charlson Comorbidity Index score; SpO2/FIO2, Pulse oximetry saturation to fraction of inspired oxygen ratio; CR, Chest radiography; ICU, Intensive care unit; Q1, first quartile; Q3, third quartile; S/CO, absorbance/cut-off; IL-6, Interleukin-6; NLR, Neutrophil-to-Lymphocyte Ratio. Summary statistics are provided as medians with interquartile ranges or numbers with percentages as appropriate.

No differences were found between groups in the Ct values on admission (E-gene, 29.1 [26.5-32.8] vs 29.7 [26.2-34.3]; p=0.909), time to first negative RT-PCR results (8 vs 13 days; P=0.237), the proportion of individuals who tested positive beyond 1-month (19% vs 35%; p=0.174) and 2-month (7.4% vs 16.8%; p=0.375) visits (Table 1), and in the Ct values at 1-month (E-gene, 35 [33.0-36.0] vs 36 [31.5-37.5]; p=0.754) or 2-month (E-gene, 38.5 [38.2-38.8] vs 38.0 [37.0-39.0]; p=1.000) visits.

There was a weaker antibody response against SARS-CoV-2 in the highest CSQ scores group, consisting of a longer time to either S-IgG or N-IgG seroconversion from illness onset (median 16 vs 11 days; p=0.012), a flatter slope (+0.4 [0-0.5] vs +1.0 [0.5-3.2] change in S/CO per day; p<0.001) until reaching peak levels, and lower peak S-IgG value (4.1 vs 6.4 S/CO; p=0.002) (Table 1 and Fig. 3a-b). Accordingly, fewer patients within the highest CSQ scores showed seroconversion at 2 months for S-IgG and for N-IgG (55.6% vs 86.7%; p=0.002).

**Figure 3.**
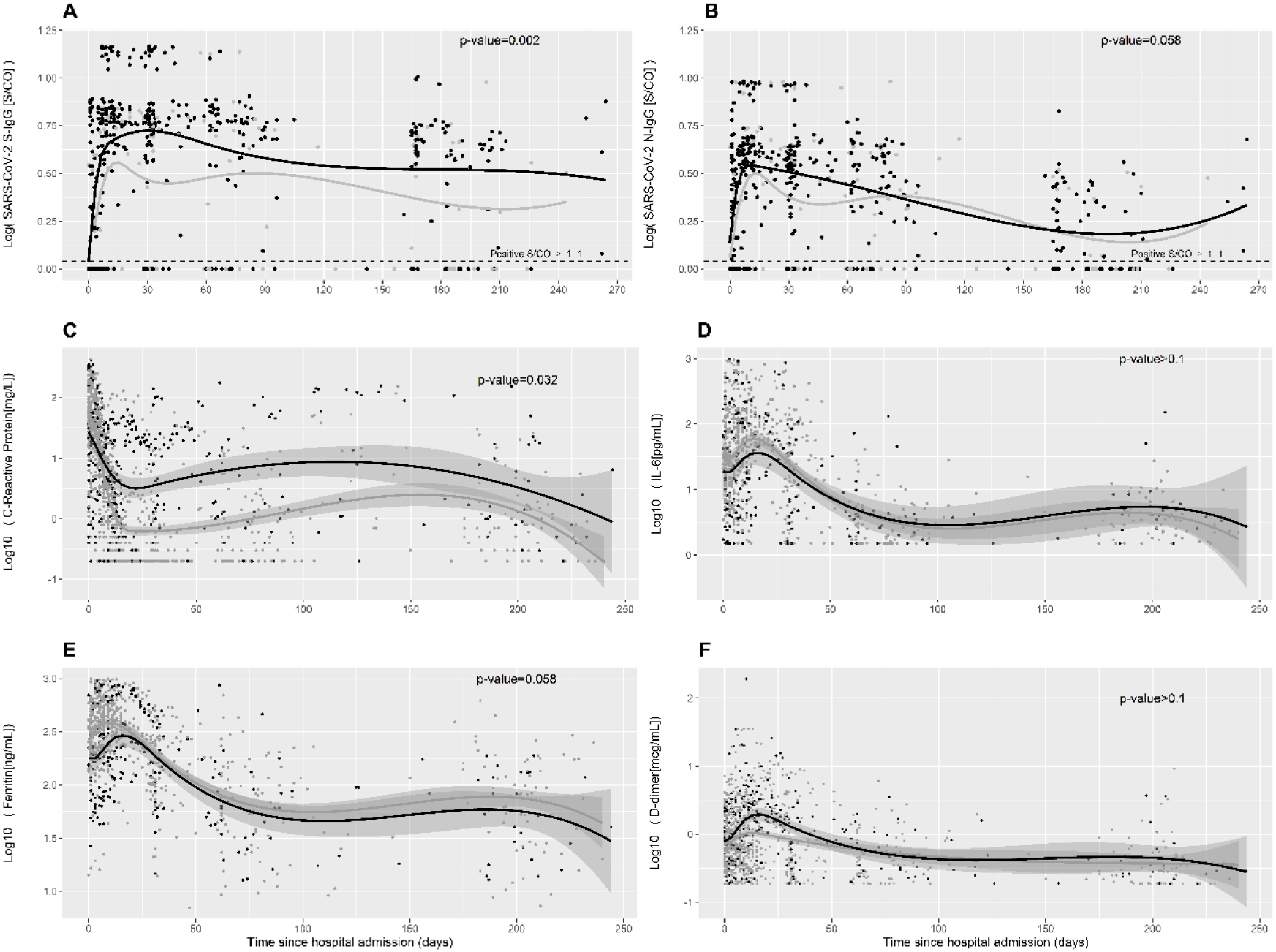
Temporal changes in SARS-CoV-2 IgG antibodies and serum inflammatory biomarkers during follow-up by symptomatic status. Serum levels of S-IgG (panel A), N-IgG (panel B), C-reactive protein (Panel C), interleukin-6 (Panel D), ferritin (Panel E) and D-dimer (Panel F) during follow up. Black lines and dots represent individuals with the highest scores in COVID-19 symptoms questionnaire; grey lines and dots represent all other patients filling the questionnaire. Abbreviations: S-IgG, IgG antibody against the SARS-CoV-2 surface S1 domain of the spike protein; N-IgG, IgG antibody against the SARS-CoV-2 internal nucleocapsid protein; S/CO, absorbance/cut-off. *P*-value for the comparison between groups according to the score in COVID-19 symptoms questionnaire.

Receiver operating characteristic (ROC) curve analysis showed that a SARS-CoV-2 S-IgG value below 5.4 S/CO at 1-month post-discharge predicted the highest CSQ scores at 2 months with an AUC [CI 95%] of 0.66 [0.54-0.79] (Fig. S-3).

A differential profile in the dynamics of CRP levels was observed according to group, with a trend to lower levels at baseline in individuals with highest CSQ scores, and subsequent inversion in the trend at 1-month and 2-month visits (Fig. 3c-f and Table 1). Baseline ferritin levels were higher in the group with highest CSQ scores. No differences were found in other inflammatory biomarkers among groups.

In the multivariate logistic regression analysis including age, sex, Charlson comorbidity index, WHO severity ordinal scale score, peak S-IgG values, testing positive for SARS-CoV-2 RT-PCR at 1-month visit and tocilizumab use, we found that a lower peak in SARS-CoV-2 S-IgG value (OR [95% CI] 0.80 [0.66-0.94]), and a higher WHO severity scale score (2.57 [1.20-5.86] per point increase) were independent predictors of the highest CSQ scores at 2-months after discharge (Fig. 4a). Sensitivity analyses with the outcomes median-CSQ score or any symptom-CSQ showed similar trends (Fig. S-4a).

**Figure 4.**
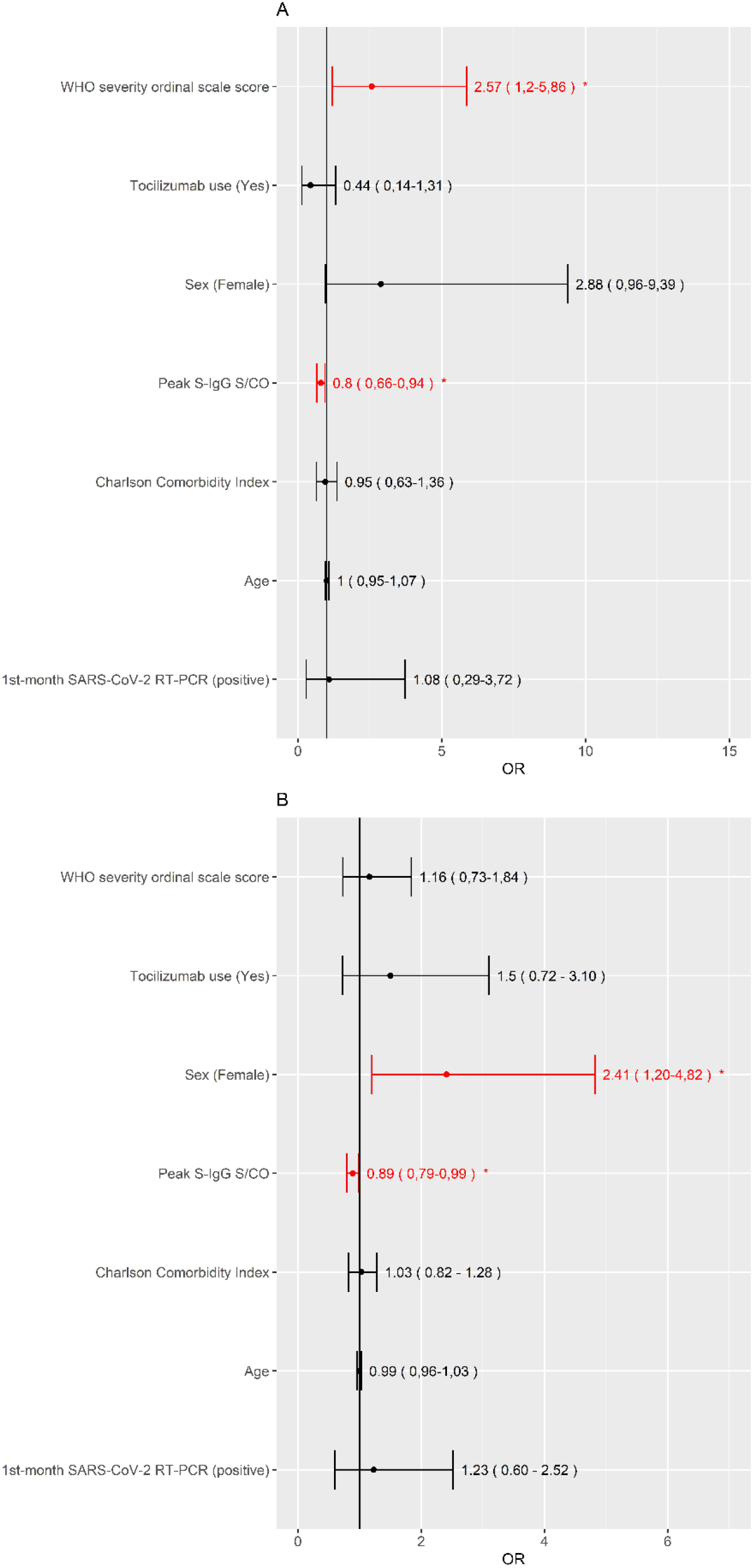
Predictors of the highest scores in COVID-19 symptoms questionnaire in multivariate regression logistic model at two-month and six-month follow-up. Panel A represents 2-month, and panel B 6-month follow-up. Abbreviations: WHO, World Health Organization; S-IgG, IgG antibody against the SARS-CoV-2 surface S1 domain of the spike protein; S/CO, absorbance/cut-off; RT-PCR, reverse transcriptase-polymerase chain reaction. \**P* value <0.05.

### Characterization and predictors of long-term persistence of symptoms

One hundred and sixteen individuals, including 85 who also had completed the survey at two months, voluntarily agreed to fill out the CSQ at the 6-month visit. The most frequent symptoms reported by patients with the highest CSQ scores were fatigue (10.3%), myalgia (6.9%), dyspnoea (4.3%), cough (4.3%) and nasal congestion (4.3%).

Patients with the highest CSQ scores showed lower CRP levels on admission and a trend to lower peak S-IgG value (Table 1).

In the multivariate logistic regression analysis including the same variables as in 2-month visit, a lower peak in SARS-CoV-2 S-IgG (OR [95% CI] 0.89 [0.79-0.99]) and female sex (2.41 [1.20-4.82] were predictors of the highest CSQ scores at the 6-month visit (Fig. 4b). Sensitivity analyses with median-CSQ score and any symptom-CSQ showed similar trends (Fig. S-4b).

## DISCUSSION

In this closely followed-up cohort evaluating the clinical, serological and virological outcomes of patients admitted to hospital for COVID-19 during a 6-month period, we observed a high frequency of clinical events following hospital discharge. Most of these events were related to patients’ underlying comorbidities, secondary bacterial infections and recurrence or complications associated with COVID-19, including thrombohemorrhagic events. A proportion of patients reported persistent symptoms in the medium (2-month) and half of them in the long-term (6-month). Chronic persistence of symptoms was associated with a distinct immunological and clinical profile, which differed according to duration of symptoms. Patients with the highest CSQ scores both in the mid-term and long-term exhibited a weaker initial antibody response and lower inflammation during the acute phase of the disease. Lasting symptoms in the mid-term were associated with persistent inflammation at months 1 and 2 after discharge and higher WHO severity score. Female sex predicted long-term complaints.

To the best of our knowledge, this is the first longitudinal study evaluating the clinical, immunological and virological outcomes and their interdependencies during a 6-month period in COVID-19 patients discharged from hospital. Patients included in this longitudinal study were comprehensively monitored. From the virological perspective, after a median duration of viral shedding of around two weeks, we found evidence of prolonged viral replication, as shown by persistent RT-PCR test positivity (or re-positivity) two months after discharge in 12% of the patients, but only four cases had a RT-PCR positive in the very late phases of follow-up. Interestingly, most patients with late RT-PCR positivity had high Ct values, and none of them fulfilled the CDC criteria for potential reinfection [8]. Moreover, genotypic sequencing confirmed the same clade in all analyzed samples, suggesting that they represented persistent infection/reactivation.

Our study is one with the largest follow-up to assess the kinetics of the immune humoral response over time. S-IgG antibodies reached higher titers, followed by a gradual decline of both S-IgG and N-IgG. Seroreversion was observed for N-IgG titers in more than one fourth of patients at 6 months after discharge, but it was infrequent with the potentially neutralizing S-IgG antibodies, which implies a durable antibody response in patients with severe COVID requiring hospitalization, in contrast to the rapid waning of antibodies reported in patients with mild COVID [9-11]. Severity of disease has actually been associated with the magnitude and duration of the antibody response [12].

While long-COVID is increasingly being described after acute infection, the pathogenesis and duration of this multifaceted syndrome remain unknown. Data about long-COVID with such a lengthy follow-up are very scarce [13]. Our data suggest an involvement of the antibody response in the occurrence of long-COVID. The highest CSQ scores in the medium and long term were associated with a lower peak of S-IgG, and in patients with mid-term symptoms a delayed antibody response was additionally observed, the latter also showing to be a predictor of the highest CSQ scores in multivariate analysis (data not shown). Experimental studies have demonstrated that, in addition to T-cell response, IgG and neutralizing antibodies are crucial to control viral infections [14,15]. Besides the antimicrobial activity, antibodies contribute to modulate the inflammatory response via the Fc-gamma receptor, toll-like receptors and activation of the complement, all of them inducing the secretion or repression of various pro-inflammatory and anti-inflammatory mediators [16-19]. Both the antiviral and the immunomodulatory effects of antibodies might have been involved in the enhanced recovery of patients with higher antibody levels in our cohort. The protective effects of immunoglobulins are actually the basis for the use of convalescent plasma [20,21] or neutralizing monoclonal antibodies against SARS-CoV-2 spike protein [22] to treat patients with acute COVID-19. We calculated the cut-point for S-IgG at one month that predicted the ulterior persistence of highest CSQ scores. Accordingly, the use of monoclonal antibodies or boosting the antibody response with vaccination might be potential strategies to prevent long-COVID in patients with antibody levels under these cut-offs [22]. This hypothesis warrants further research.

Post-COVID syndrome was associated with additional distinctive innate and adaptive immune traits, consisting of a weaker initial inflammatory response, as shown by lower baseline levels of CRP and ferritin. This is aligned with the poorer antibody response observed in this subset of patients, and supports the correlation described between antibody levels and inflammation biomarkers [23]. Potent innate and adaptive immune responses during acute infection might potentially lead to a more efficient control of illness and lower residual inflammation and incidence of subsequent sequelae. Interestingly, patients with mid-term lasting symptoms showed persistent residual inflammation. Elevated markers of inflammation and autoimmunity have been reported in post-infectious syndromes linked with Chikungunya or Epstein-Barr viruses, or in myalgic encephalomyelitis [24-27].

Besides the immune response, demographic and clinical factors were found to be predictors of long-COVID. Long-term persistent symptoms were associated with female sex, a finding also supported by other studies [13,28]. Higher severity of disease showed to be a predictor of mid-term persistent symptoms. Severe disease would be expected to induce greater organ injury, which might be associated with higher incidence of sequelae, as supported by the higher prevalence of persistent symptoms found in patients with COVID-19 admitted to the ICU [13,29]. Although post-discharge complaints have also been described in patients with milder forms of COVID, critically-ill patients have more frequently shown subjacent structural causes contributing to explain the symptoms [30], and up to 80% of them develop physical, cognitive or mental health impairments that persist beyond hospital discharge [31]. Differences in patients’ characteristics and in the intensity of symptoms might have contributed to the higher degree of reversibility potentially occurring in our patients, in whom severity of disease did not show to be a predictor of long-term high-score symptoms compared to the results of Huang et al [13].

The sample size is a limitation of the study. Long-COVID is a heterogeneous syndrome and the associations found might differ according to the nature of the symptoms analyzed. We focused on subjects reporting the highest scores in the questionnaire to identify those symptoms that were more likely to be meaningful to patients and avoid overrepresentation of long-COVID cases. However, sensitivity analyses including patients with any symptom, or those with a score above the median, showed similar results. Strengths are the longitudinal design with consecutive sampling, close monitoring and thorough investigations conducted in the patients.

In conclusion, long-term follow-up of patients hospitalized for COVID shows a high frequency of clinical events after hospitalization, a durable antibody response of S-IgG, and frequent RT-PCR test positivity/re-positivity occurring beyond two months after acute infection, but with no evidence of reinfection. Persistent symptoms are common in the medium and long term. Gender, severity of illness and the immune response are associated with long-COVID, but with different implication according to the temporality of symptoms. The antibody response predicts both mid-term and long-term clinical outcomes, and consequently the use of monoclonal antibodies or boosting the antibody response with vaccination might be potential strategies to prevent long-COVID.

## Data Availability

Data is not publicly available due to IRB restrictions.

