## Supplemental material for "Long-term clinical, virological and immunological outcomes in patients hospitalized for COVID-19: antibody response predicts long COVID"

### Supplementary material

#### A) Clinical and sero-virological supplementary material

##### Table S-1. Baseline characteristics and clinical events occurring during the six months follow-up.

| Variable | | |
|---|---|---|
| Patients, no. | 146 | (100) |
| Male sex | 88 | (60.3) |
| Age, years | 64 | (54-76) |
| Current smoking | 12 | (8.8) |
| **Comorbidity** | | |
| Any comorbidity* | 106 | (72.6) |
| CCI, median (Q1, Q3) points | 3 | (1-5) |
| Cardiovascular disease | 31 | (21.2) |
| Hypertension | 32 | (21.9) |
| Diabetes | 33 | (22.6) |
| Chronic obstructive lung disease | 8 | (5.5) |
| Symptoms duration, median (Q1, Q3) | 7 | (3-10.5) |
| No. of symptoms on admission | 3 | (2-4) |
| Maximum WHO severity score | 4 | (4-4) |
| SpO2/FIO2 on admission | 350 | (339.3-452.4) |
| Bilateral lung infiltrates in CR | 122 | (89.1) |
| Length of hospital stay, days | 11 | (8-15) |
| **Treatments** | | |
| Dexamethasone/Methylprednisolone | 23 | (15.8) |
| Tocilizumab | 73 | (50) |
| **Sero-virological features** | | |
| SARS-CoV-2 nasopharynx viral quantification | 2227 | (149 - 23382) |
| SARS-CoV-2 Cycle threshold | 34.7 | (27-40) |
| Time to viral clearance | 13 | (2.2-33.8) |
| RT-PCR SARS-CoV-2 positive, n (%) | | |
| ≥1 month | 40 | (27.4) |
| ≥2 months | 15 | (11.8) |
| ≥6 months | 4 | (3.0) |
| Time to Seroconversion, days | 12 | (8-15) |
| SARS-CoV-2 IgG-S | | |
| Seropositivity at 1 month, n (%) | 93 | (70.5) |
| Seropositivity at 2 month, n (%) | 96 | (70.6) |
| Seropositivity at 6 month, n (%) | 99 | (71.2) |
| Peak serum concentration (S/CO) | 5.9 | (0.3-7.1) |
| Trough serum concentration (s/CO) | 3.9 | (2.1-4.7) |
| SARS-CoV-2 IgG-N | | |
| Seropositivity at 1 month, n (%) | 94 | (71.2) |
| Seropositivity at 2 month, n (%) | 96 | (70.6) |
| Seropositivity at 6 month, n (%) | 98 | (70.5) |
| Peak serum concentration (S/CO) | 4.1 | (0.3-4.9) |
| Trough serum concentration (s/CO) | 2.2 | (1.5-2.8) |
| **Clinical events during FU** | | |
| Deaths | 8 | (5.48) |
| Persistence of any symptoms* | | |
| At 2 months | 27 | (25.9) |
| At 6 months | 28 | (24.1) |
| **Inflammatory biomarkers** | | |
| Serum C-Reactive-protein, mg/L | | |
| On admission | 39.1 | (15.2-89.9) |
| 1 month | 0.3 | (0.2-2.1) |
| 2 months | 0.5 | (0.2-4.2) |
| 6 months | 1 | (0.4-5.1) |
| Serum IL-6, pg/mL | | |
| On admission | 25.1 | (10.9-73.6) |
| 1 month | 15.6 | (3.5-54.9) |
| 2 months | 5.2 | (2.2-20.5) |
| 6 months | 3 | (1.8-5.1) |
| Serum Ferritin, ng/mL | | |
| On admission | 354 | (174-579) |
| 1 month | 181 | (90-346) |
| 2 months | 121 | (52-208) |
| 6 months | 75 | (31-134) |
| Serum D-dimer, mcg/mL | | |
| On admission | 0.7 | (0.4-1.7) |
| 1 month | 0.4 | (0.2-1.3) |
| 2 months | 0.4 | (0.3-0.8) |
| 6 months | 0.4 | (0.2-0.7) |
| NLR | | |
| On admission | 4.5 | (2.9-6.5) |
| 1 month | 1.7 | (1.2-2.7) |
| 2 months | 1.8 | (1.3-2.6) |
| 6 months | 2.0 | (1.4-2.9) |

*Individuals with a score equal or more than the third quartile in any of the CSQ items. CSQ, COVID-19 symptoms questionnaire; CCI, Charlson Comorbidity Index score; CR, chest radiograph; FU, follow-up; ICU, Intensive Care Unit; Q1, first quartile; Q3, third quartile; IL-6, Interleukin-6; SpO2, pulse oximetry saturation; FiO2, fraction of inspired oxygen; S/CO, absorbance/cut-off; NLR, Neutrophil-to-Lymphocyte Ratio. Summary statistics are provided as medians with interquartile ranges or numbers with percentages as appropriate. Results of RT-PCR SARS-CoV-2 were available in 146, 127 and 134 patients at 1, 2 and 6 months visits.

**Table S-2 Causes of in-hospital and after discharge deaths, hospital readmissions and ED attendances in the 162 COVID-19 patients admitted to Hospital General Universitario de Elche between March 1, and June 30, 2020.**

| | Events | | | |
|---|---|---|---|---|
| | Deaths | | Hospital readmission[&] | ED attendances[&] |
| | In-hospital | After discharge | | |
| All causes, no. | 12 | 8 | 30/54 | 58/117 |
| Bacterial infection, no. | 6 | 4 | 5/12 | 11/27 |
| Underlying disease exacerbation, no. | 4 | 3 | 13/23 | 16/31 |
| Thrombohemorrhagic events, no. | 2 | 1 | 9/9 | 9/9 |
| COVID-19 related, no. | - | 0 | 6/8 | 14/14 |
| Unknown/Others, no. | | | 2/2 | 23/36 |

& Each of these two columns show the number of patients (left) and the number of events (right). Some patients had more than one event. ED, Emergency department.

**Figure S-1. Flow chart of patients with COVID-19 admitted to Hospital General Universitario de Elche between March 1, and June 30, 2020.**

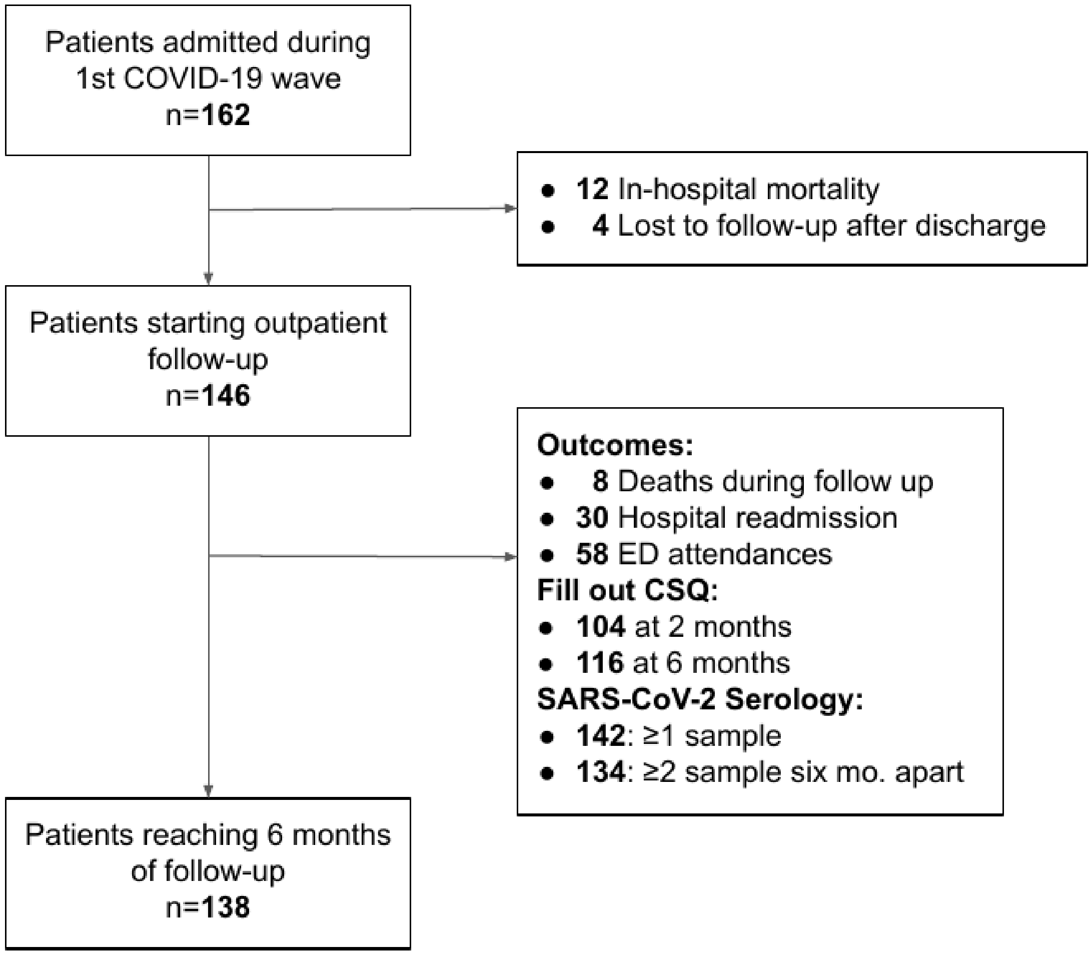


ED, Emergency Department; CSQ, COVID-19 symptoms questionnaire; mo., months. Of the eight deaths, two occured before reaching two months visits.

**Figure S-3. ROC curve of anti-SARS-CoV-2 spike IgG S/CO values at 1st month in predicting two months UQ-CSQ.**

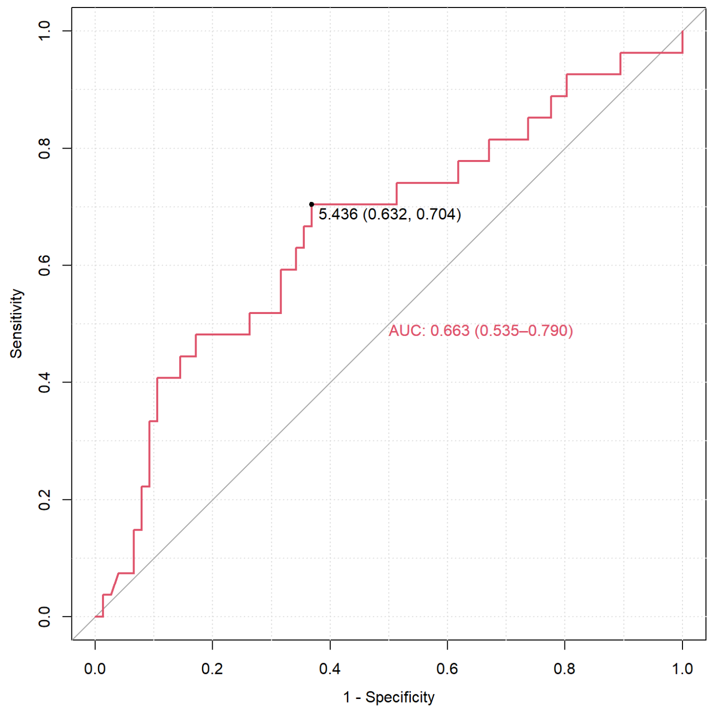


UQ-CSQ, Upper Quartile-CSQ: individuals with a score equal or more than the third quartile in any of the CSQ items. CSQ, COVID-19 symptoms questionnaire. ROC, Receiver operating characteristics; AUC, area under the curve; S/CO, absorbance/cut-off.

**Figure S-4. Sensitivity analyses for predictors, in multivariate regression logistic model, of median-CSQ and any symptom-CSQ at two (panels A and B, respectively) and six (panel C and D, respectively) months of follow-up.**

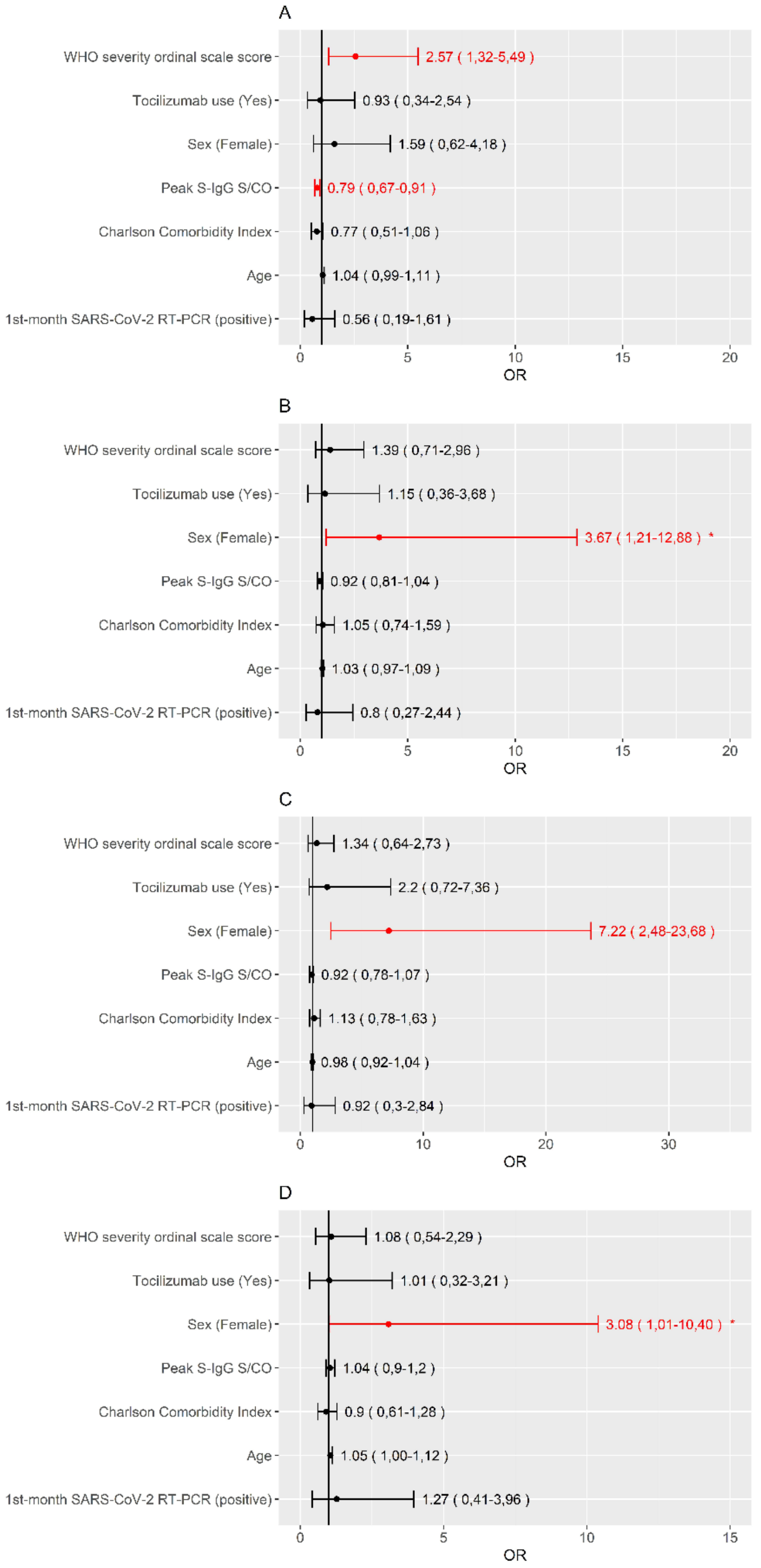


Median-CSQ, individuals with a score equal or more than the median in any of the CSQ items. Any symptom-CSQ, individuals with a score at least one point in any of the CSQ items. WHO, World Health Organization; S-IgG, IgG antibody against the SARS-CoV-2 surface S1 domain of the spike protein; S/CO, absorbance/cut-off; RT-PCR, reverse transcriptase-polymerase chain reaction. *$P$ value <0.05. **$P$ value <0.01.

#### B) SARS-CoV-2 sequencing supplementary material

##### Genome Sequencing

Genome sequencing of SARS-COV2 was performed on stored RNA eluates from NPS samples following ARTIC amplicon sequencing protocol for MinION version V3 (https://www.protocols.io/view/ncov-2019-sequencing-protocol-bbmuik6w).

The sequencing protocol starts with a cDNA synthesis of the genome of the virus followed by a PCR that generates 400 bps amplicons in a tiled fashion across the whole SARS-COV2 genome. After samples library synthesis, the samples were sequenced with the MinION nanopore sequencer (Oxford Nanopore Technologies, Oxford, UK) using the Ligation sequencing kit with the Native barcoding expansion labeling each sample with a different barcode which allowed their multiplex sequencing. The sequencing runs were performed using the flow cell FLO-MN106D and extended until samples got 50.000 high quality reads. Sequencing basecalling was made using the nanopore guppy basecaller with the profile high accuracy base calling.

##### Bioinformatics analysis

Downstream analysis was performed following ARTIC nCOV-2019 bioinformatics protocol (https://artic.network/ncov-2019/ncov2019-bioinformatics-sop.html), a protocol which takes the output from the sequencing protocol to get the consensus genome SARS-COV2 sequences, the protocol follows the basecalling step and make a de-multiplexing sample sequences followed by a sampling mapping step polishing and a sequence consensus generation.

Phylogenetic analysis of paired specimens was done using webserver Nexstrain (https://nextstrain.org/), with the SARS-COV2 database Nextclade (https://clades.nextstrain.org/) to identify the clade, mutation calling and phylogenetic placement of the SARS-COV2 genomes (GISAID genoma data).

**Figure S-2.** Identification of the samples in the phylogenetic tree of reference genome sequences of SARS-CoV-2 from GISAID database.

Individual #95: Samples 70-MFB from March 26, 2020 and 765-MFB from September 10, 2020.
Individual #88: Samples 248-FMMS from April 7, 2020 and 722-FMMS from September 24, 2020.
Individual #8: Samples 419-IVG from March 30, 2020 and 745-IVG from September 10, 2020.

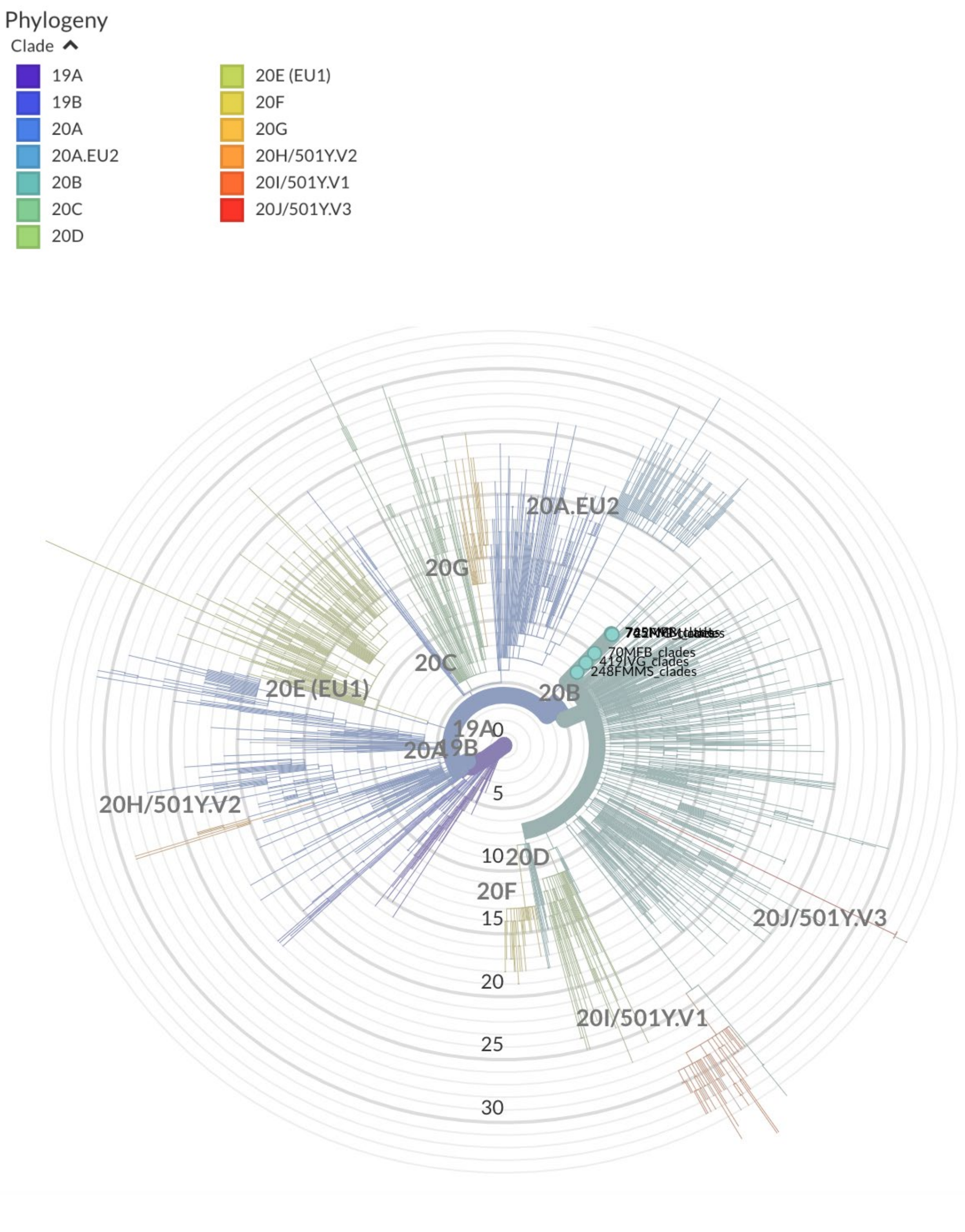

**Table S-3. Aminoacid substitutions in the samples versus GISAID reference genomes.**

| | Sample | | Aminoacid substitutions |
|---|---|---|---|
| **Individual #95** | 70-MFB (March 26, 2020) | 765-MFB (September 10, 2020) | |
| | Yes | Yes | N: R203K |
| | Yes | Yes | N: G204R |
| | Yes | Yes | N: S235F |
| | Yes | Yes | N: K374R |
| | Yes | Yes | ORF1b: P314L |
| | Yes | Yes | S: D614G |
| **Individual #88** | 248-FMMS (April 7, 2020) | 722-FMMS (September 24, 2020) | |
| | Yes | Yes | N: R203K |
| | Yes | Yes | N: G204R |
| | Yes | Yes | N: S235F |
| | Yes | Yes | N: K374R |
| | Yes | Yes | ORF1b: P314L |
| | Yes | Yes | S: D614G |
| **Individual #8** | 419-IVG (March 30, 2020) | 745-IVG (September 10, 2020) | |
| | Yes | Yes | N: R203K |
| | Yes | Yes | N: G204R |
| | Yes | Yes | N: S235F |
| | No | Yes | N: K374R |
| | Yes | Yes | ORF1b: P314L |
| | No | Yes | S: A222V |
| | Yes | Yes | S: D614G |
